# The influence of place on COVID-19 vaccine coverage in Alberta: A multilevel analysis

**DOI:** 10.1101/2022.06.15.22276467

**Authors:** Yuba Raj Paudel, Crystal Du, Shannon E. MacDonald

## Abstract

**Background:** While there is evidence of urban/rural disparities in COVID-19 vaccination coverage, there is limited data on the influence of other place-based variables.

**Methods:** In this cross-sectional study, we analyzed population-based linked administrative health data to examine vaccination coverage for 3,945,103 residents in Alberta, Canada. We used multilevel logistic regression to examine the association of vaccination coverage with various place-based variables.

**Results:** After 4 months of widely available COVID-19 vaccine, coverage varied widely between rural and urban areas (58% to 73%) and between geographic health authority zones (55.8% to 72.8%). Residents living in neighborhoods with lower COVID-19 disease incidence had the lowest vaccination coverage (63.2%), while coverage in higher incidence neighborhoods ranged from 68.3% to 71.9%. The multilevel logistic regression model indicated that residence in metro (adjusted odds ratio [aOR] 1.37; 95% CI: 1.31-1.42) and urban areas (aOR 1.11; 95% CI: 1.08-1.14) was associated with higher vaccine coverage than residence in rural areas. Similarly, residence in Edmonton, Calgary, and South health zones was associated with higher vaccine coverage compared to residence in Central zone. Higher income neighborhoods reported higher vaccine coverage than the lowest-income neighborhoods, and the highest COVID-19 risk neighborhoods reported higher vaccine coverage than the lowest risk neighborhoods (aOR 1.52; 95% CI: 1.12-2.05).

**Conclusion:** In the first four months of wider vaccine availability in Alberta, COVID-19 vaccine coverage varied according to various place-based characteristics. Vaccine distribution strategies need to consider place-based variables for program prioritization and delivery.

## Introduction

Successful control of the COVID-19 pandemic depends on collective action, including rapid and widespread vaccination.(1)It is well known that vaccine coverage levels differ by individual-based variables (e.g. age, attitudes toward vaccines), but place-based variables also play a role.(2)

Rural-urban residence is a variable commonly included in analyses of COVID-19 vaccination coverage. Studies from the USA have shown variability in vaccine coverage in rural versus urban regions. Data from April 2021 showed COVID-19 vaccine coverage among rural residents was 58.5% compared to 75.4% among urban residents(3), although individual-level characteristics were not considered. Another study found similar results, in addition to variation among rural counties.(4)

The reasons for lower vaccine uptake in rural regions are multifaceted, as seen in a number of studies form the USA. One study suggested that the disparity was linked with lower access to health care in rural counties, as well as higher vaccine hesitancy and low-risk perception.(3) Another study showed that areas with slow vaccine rollout were more likely to be the regions with high barriers related to healthcare access and resources.(1) These areas were historically under-vaccinated, had irregular healthcare-seeking behaviour, and more socio-demographic barriers.(1) Their data also showed geographic clustering of barriers, with some counties exhibiting more barriers than others. Another study showed lower educational attainment was associated with low vaccination in rural areas.(4) Further, rural counties with farming and mining dependent economies had lower coverage than rural counties with recreation-dependent economies.

While rural-urban differences for COVID-19 vaccination are well described in the US literature, there are other less-studied factors related to place that may impact coverage. Place-based factors need to be considered to better understand community-level barriers for COVID-19 vaccine uptake. In this study, we examined the association between COVID-19 vaccination status and place-related variables, including rural-urban residence, geographic health zone, neighborhood income quintile, and neighborhood COVID-19 risk quintile, while also adjusting for individual-level demographic variables (age and sex).

## Methods

### Setting

This study was conducted in Alberta, a western province of Canada with a population of 4.5 million. Alberta has universal health care insurance under which over 99% of Albertans are registered. Publicly funded COVID-19 vaccine was widely available throughout the province starting mid-May 2021 for all Albertans 12 years and older.

### Study design, and data sources

This was a population-based cross-sectional study using administrative data retrieved from the Alberta Ministry of Health data repositories. We used the Alberta Health Care Insurance Plan (AHCIP) quarterly population registry for the first quarter of 2021 to identify residents of the province. We used the Immunization and Adverse Reaction to Immunization (Imm/ARI) repository to determine COVID-19 vaccination status (as of August 31 2021). Imm/ARI includes all records of COVID-19 vaccines administered in Alberta, regardless of provider. The total number of individuals who tested positive for COVID-19 was derived using laboratory data (as of August 31 2021). We deterministically linked the databases using unique personal health numbers.

### Study Cohort

Our cohort included all Alberta residents (as of first quarter of 2021) aged 12 years and above. Albertans <12 years were excluded because they were not eligible to receive the COVID-19 vaccine during the study period. We excluded First Nations residents of Alberta (since data for First Nations communities was not consistently submitted to Imm/ARI), Lloydminster residents (since vaccines are delivered by the neighbouring province), and those who left the province or died during the study period. Appendix Figure A1 presents sample selection flow chart.

### Outcome measure

We calculated ‘COVID-19 vaccination coverage’ as the proportion of eligible Alberta residents who received at least one dose of a COVID-19 vaccine as of August 31 2021. We chose this time point to examine vaccination coverage during a time when vaccines were widely available.

### Exposure variables

#### Individual characteristics

We divided participant age into six categories, based on age categories used to prioritize COVID-19 vaccine eligibility in Alberta: 12-17 years, 18-29 years, 30-49 years, 50-64 years, 65-74 years, and 75 years and above. Biological sex at birth was categorized into male and female.

#### Place-based characteristics

Place-based variables, identified by complete 6-digit postal code, included: (1) rural-urban residence, categorized into metro and moderate metro, urban and moderate urban, and rural and remote rural, based on 2016 Canadian census data; (2) five geographic health zones of the province used for local health care program decision making (South, Calgary, Central, Edmonton, and North zones); (3) neighborhood income quintiles (Q1 indicating the lowest income neighborhood and Q5 indicating the highest income neighborhood), based on the 2016 Canadian census. One place-based variable, (4) neighborhood COVID-19 risk quintile, was identified at the Forward Sortation Area (FSA) level, derived from the first 3 digits of the postal code. Neighborhood COVID-19 risk quintile was defined based on the cumulative number of people that tested positive for COVID-19 out of the total population of the FSA (expressed as proportion/10,000 population). Risk quintiles were defined as: >700 (risk quintile 1, includes 8 FSAs), 550-700 (risk quintile 2, includes 23 FSAs), 450-550 (risk quintile 3, includes 32 FSAs), 350-450 (risk quintile 3, includes 57 FSAs), <350 (risk quintile 5, includes 30 FSAs).

### Statistical analysis

We measured vaccination coverage by place-related variables: urban/rural place of residence, geographic health zone, neighbourhood income quintile, and neighborhood COVID-19 risk quintile. We used multilevel logistic regression to estimate effect sizes, adjusting for individual-level variables (sex and age), while accounting for clustering within each FSA. Before running multilevel logistic regression, we tested for multicollinearity among predictors, and no strong correlation was observed.

The final model included variables at the individual level (variables defining characteristics of the individuals [age categories and sex]; place-based variables defined at the postal code [rural-urban place of residence, geographic health zone, neighbourhood income quintile], and a variable at the FSA level [neighborhood COVID-19 risk quintile]. To calculate intra-class correlation, we assumed that the dichotomous outcome variable (receipt of at least one dose of vaccine) came from a latent variable (continuous) with a level-1 residual which follows a logistic distribution having a mean equal to 0 and a variance equal to 3.29.(5) Intra-class correlation (ICC) is a measure of correlation in an outcome variable among individuals living within a same unit (i.e. school, neighborhood). In other words, ICC indicates the amount of variation in vaccination coverage that is accounted for by variation between groups (between FSA variations).(6) An ICC value of 0 indicates that all variability in vaccination lies within the FSA. Whereas, an ICC value of 1 indicates all variability in vaccine coverage lies between FSAs (individuals within an FSA have the same vaccine coverage).(7) Literature suggests that chances of type I error (showing a significant effect, when in fact when no effect exists) increases with increasing value of ICC.(7) Therefore, hierarchically structured data need to be analysed using multilevel models.

We then combined information on vaccination coverage and neighborhood COVID-19 risk quintiles to define and map FSA categories on a map of the province, using QGIS software.

We performed statistical analysis using SAS 9.4 (SAS Institute Inc., Cary, NC) with a statistical significance set at p<0.05. The University of Alberta Health Research Ethics Board granted ethical approval for this study.

## Results

### Cohort characteristics

Of the total population included in this analysis (n=3,945,103), 49.6% were female. The majority (70%) lived in metropolitan areas (Table 1). By geographic health zones, Calgary (40%) and Edmonton Zone (32.7%) contained the majority of the population. Less than 10% of the total population lived in neighborhoods with the highest COVID-19 risk quintile; most of the population lived in neighborhoods with medium to low medium risk quintiles (60%).

**Table 1.** COVID-19 vaccination coverage by individual and place-based characteristics, as of August 31 2021 in Alberta, Canada (N=3,945,103)

| Variables | Vaccinated<br>(≥ one dose)<br>% (n) | Unvaccinated<br>(no dose)<br>% (n) | p value |
| --- | --- | --- | --- |
| Age category |  |  |  |
| 12-17 years | 62.9 (205,240) | 37.1(120,827) | <0.0001 |
| 18-29 years | 62.9 (424,326) | 37.1 (249,843) |  |
| 30-49 years | 64.4 (918,233) | 35.6 (507,803) |  |
| 50-64 years | 74.5 (642,084) | 25.5 (219,683) |  |
| 65 to 74 years | 81.1 (323,157) | 18.9 (75,195) |  |
| 75 years and above | 82.0 (212,087) | 18.0 (46,625) |  |
| Sex |  |  |  |
| Females | 71.3 (1,396,547) | 28.7 (561,962) | <0.0001 |
| Males | 66.9 (1,328,580) | 33.1 (658,014) |  |
| Place of residence |  |  |  |
| Metro | 73.1 (2,010,401) | 26.9 (738732) | <0.0001 |
| Urban | 62.4 (299,103) | 37.6 (179973) |  |
| Rural | 58.0 (415,623) | 42.0 (301271) |  |
| Geographic health zone |  |  |  |
| South | 63.4 (171,150) | 36.6 (98,737) | <0.0001 |
| Calgary | 72.7 (1,148,512) | 27.3 (432,228) |  |
| Central | 59.9 (241,388) | 40.1 (161,526) |  |
| Edmonton | 72.8 (940,818) | 27.2 (350,869) |  |
| North | 55.8 (223,259) | 44.2 (176,616) |  |
| Neighborhood income quintile |  |  |  |
| Highest income (Q5) | 72.7 (595,519) | 27.3 (223,305) | <0.0001 |
| Q4 | 70.8 (586,582) | 29.2 (242,565) |  |
| Q3 | 69.6 (552,301) | 30.4 (240,963) |  |
| Q2 | 67.4 (530,707) | 32.6 (257,121) |  |
| Neighborhood COVID-19 risk quintile |  |  |  |
| Highest risk (R1) | 71.9 (237,315) | 28.1 (92,763) | <0.0001 |
| R2 | 68.3 (377,240) | 31.7 (175,481) |  |
| R3 | 71.5 (664,873) | 28.5 (264,804) |  |
| R4 | 70.0 (1,004,100) | 30.0 (429,697) |  |
| Lowest risk (R5) | 63.2 (441,599) | 36.8 (257,231) |  |

### Vaccination coverage by place-related variables

The highest vaccination coverage was among people living in metro areas (73.1%), followed by those living in urban and moderate urban areas (62.4%), with the lowest coverage in rural areas (58.0%). By geographic health zones, Edmonton (72.8%) and Calgary (72.6%) had higher coverage compared to North (55.8%), Central (59.9%), and South (63.4%) zones. The highest income neighbourhoods (Q5) had the highest vaccination coverage (72.7%), while the lowest income neighbourhoods (Q1) had the lowest coverage (64.2%). People living in neighborhoods with lowest COVID-19 risk quintile had the lowest vaccination coverage (63.2%). Other neighborhood risk quintiles reported a similar vaccination coverage to each other ranging from 68.3% to 71.9%.

### Multilevel logistic regression

ICC from the unconditional model was 0.05 (ICC ranges from 0 to 1) indicating that there was a little variation in vaccination coverage between FSAs. The adjusted ORs from the final multilevel model (see Table 2 and Appendix Table A1) showed that participants living in metro areas (aOR 1.37; 95% CI: 1.31-1.42) and urban areas (aOR 1.11; 95% CI: 1.08-1.14) had higher vaccination coverage compared to rural residents. People living in Calgary zone (aOR 1.26; 95% CI: 1.21-1.31), South zone (aOR 1.17; 95% CI: 1.12-1.22), and Edmonton zone (aOR 1.11; 95% CI: 1.05-1.18) had higher vaccination coverage compared to Central zone. People living in North zone (aOR-0.82; 95% CI: 0.77-0.86) had lower vaccine coverage than Central zone. People residing in the highest income neighborhoods were nearly twice as likely to get vaccinated than those living in the lowest income neighborhood (aOR 1.76; 95% CI: 1.60-1.93). Neighborhoods in middle-income quintiles (Q2, Q3, Q4) also had higher vaccination coverage than the lowest income quintile neighborhoods. People living in neighborhoods with the highest COVID-19 risk had higher vaccination coverage compared to those in the lowest risk neighborhoods (aOR 1.52; 95% CI: 1.12-2.05). Although other risk quintiles had a higher crude vaccination coverage than the lowest income quintile neighborhoods, the effect disappeared in the multivariable model (Table 2 and Appendix Table A1).

**Table 2:** Factors associated with receipt of one dose of COVID-19 vaccine in Alberta, Canada.

| **Variables** | **Unadjusted OR**<br>**(95% CI)** | **Adjusted OR \***<br>**(95% CI)** |
| --- | --- | --- |
| **Age categories** |  |  |
| 12-17 years | Ref | Ref |
| 18-29 years | 1.00 (0.99-1.01) | 1.01 (0.89-1.14) |
| 30-49 years | 1.07 (1.06-1.07) | 1.04 (0.92-1.18) |
| 50-64 years | 1.72 (1.71-1.74) | 1.77 (1.56-2.01) |
| 65 to 74 years | 2.53 (2.50-2.56) | 2.66 (2.35-3.02) |
| 75 years and above | 2.68 (2.65-2.71) | 2.88 (2.54-3.26) |
| **Sex** |  |  |
| Male | Ref | Ref |
| Female | 1.23 (1.23-1.24) | 1.21 (1.20-1.23) |
| **Income Quintile** |  |  |
| Q1 (Lowest income) | Ref | Ref |
| Q2 | 1.15 (1.14-1.16) | 1.22 (1.11-1.33) |
| Q3 | 1.28(1.27-1.28) | 1.37 (1.25-1.49) |
| Q4 | 1.35 (1.34-1.36) | 1.50 (1.37-1.64) |
| Q5 (Highest income) | 1.48 (1.47-1.49) | 1.76 (1.60-1.93) |
| **Geographic health zone** |  |  |
| Central zone | Ref | Ref |
| South zone | 1.16 (1.15-1.17) | 1.17 (1.12 -1.22) |
| Calgary zone | 1.78 (1.77-1.79) | 1.26 (1.21-1.31) |
| Edmonton zone | 1.79 (1.78-1.81) | 1.11 (1.05-1.18) |
| North zone | 0.85 (0.84-0.85) | 0.82 (0.77-0.86) |
| **Place of residence** |  |  |
| Rural | Ref | Ref |
| Metro | 1.97 (1.96-1.98) | 1.37 (1.31-1.42) |
| Urban | 1.21 (1.20-1.21) | 1.11 (1.08-1.14) |
| **Neighborhood COVID-19 risk level** |  |  |
| R5 (Lowest risk) | Ref | Ref |
| R1 (Highest risk) | 1.49 (1.48-1.50) | 1.52 (1.12-2.05) |
| R2 (Higher risk) | 1.25 (1.24-1.26) | 1.14 (0.93-1.39) |
| R3 (Medium risk) | 1.46 (1.45-1.47) | 1.15 (0.96-1.38) |
| R4 (Lower risk) | 1.36 (1.35-1.37) | 1.12 (0.96-1.32) |

\*Adjusted OR based on a multilevel logistic regression model

Six categories of FSAs were mapped (Fig.1), representing the following combination of vaccination coverage and neighborhood COVID-19 risk quintiles:

**Fig.1.**
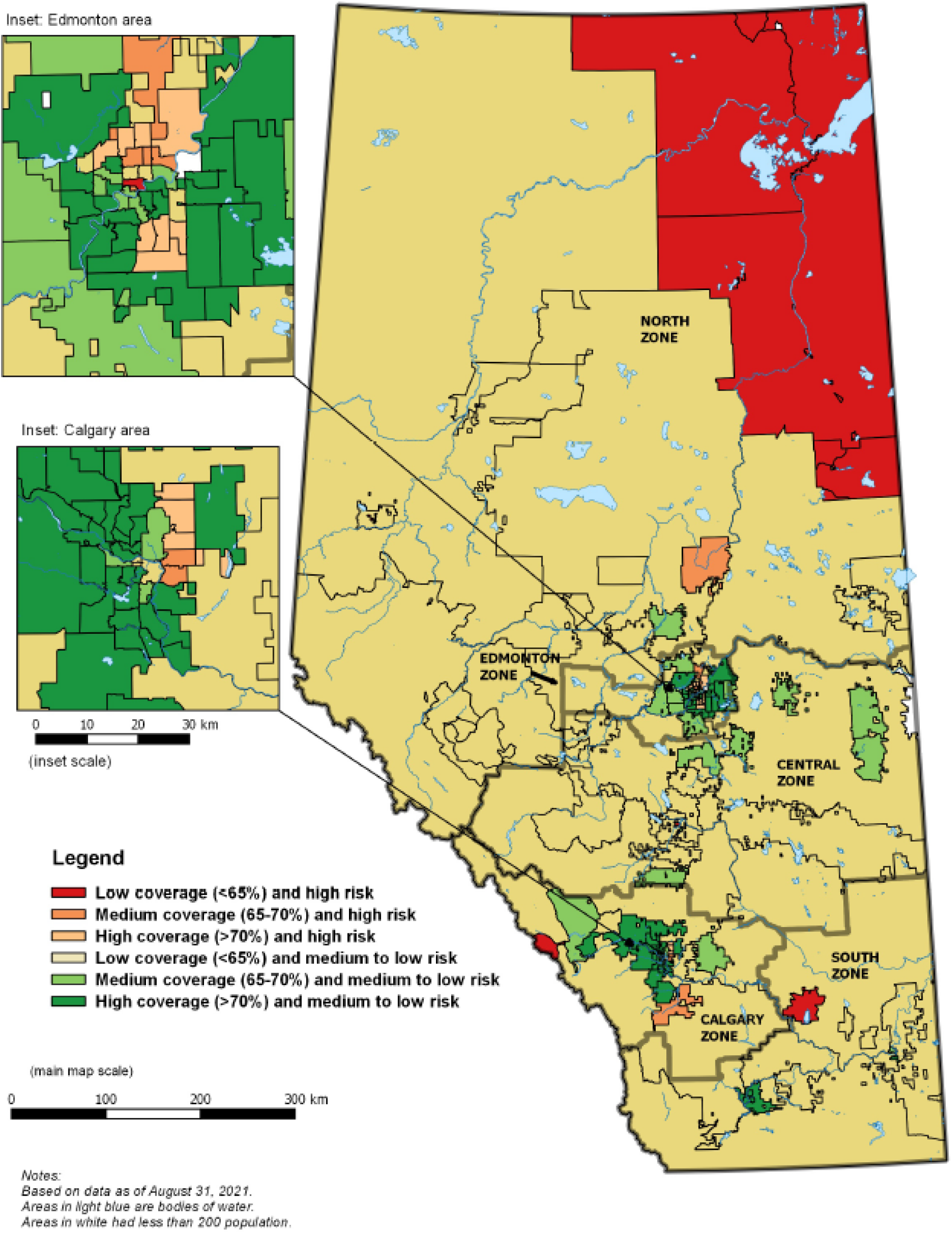
Map of Alberta forward sortation areas (FSAs) showing COVID-19 vaccination coverage and neighborhood risk quintile.

1. Low coverage (<65%) and high risk (risk quintile 1 or 2)-includes 10 FSAs
2. Medium coverage (65-70% coverage) and high risk (risk quintile 1 or 2) - includes 8 FSAs
3. High coverage (>70% coverage) and high risk (risk quintile 1 or 2) - includes 13 FSAs
4. Low coverage (<65%) and medium to low risk (risk quintile 3 to 5) - includes 38 FSAs
5. Medium coverage (65-70% coverage) and medium to low risk (risk quintile 3 to 5)-includes 27 FSAs
6. High coverage (>70% coverage) and medium to low risk (risk quintile 3 to 5)-includes 54 FSAs

Mapping vaccination coverage by neighborhood risk quintile indicated that the majority of category 6 FSAs (high coverage and medium to low risk) were located in three cities (Calgary, Edmonton and Lethbridge). The Northeast part of Calgary and Edmonton included category 2 and 3 FSAs, defined as high risk and high to medium coverage. Category 5 FSAs, defined as medium coverage and medium to low risk, were in the outskirts of these three cities. Most category 4 FSAs, defined as low coverage and medium to low risk, were in the rural areas of each zone. Category 1 FSAs, defined as low coverage and high risk, were in industrial zones of North zone (Fort McMurray, Fort McKay), South zone (Brooks), and Calgary zone (Banff).

## Discussion

### Summary

We found that place-based variables had a significant association with COVID-19 vaccination coverage after adjusting for age categories and sex. Multilevel logistic regression analysis revealed that there was higher vaccine coverage among people living in metro and urban areas compared to those in rural areas; those in higher income neighborhoods compared to poorest lowest income neighborhoods; and those in South zone, Calgary, and Edmonton compared to those living in Central zone, while those in North zone had lower coverage. People living in the highest COVID-19 risk neighborhoods had higher vaccination coverage than those living in the lowest risk neighborhoods.

### Interpretation

Our analysis showed that vaccination coverage in Alberta, as of August 2021, ranged from 58% in rural areas to 73% in large metropolitan regions, including Calgary and Edmonton. This is consistent with US findings, where vaccination coverage was consistently higher in urban centres than in neighbouring rural regions.(3) Decreased access to vaccine appointments, and challenges with vaccine supply in rural Alberta during initial stages of vaccine roll-out might have contributed to low vaccine uptake. Reportedly, vaccine supply in rural Alberta was not consistent, making management of appointments challenging, often resulting in cancelled appointments.(8) Correspondingly, the North, South, and Central health zones, which consist mainly of rural regions, reported lower vaccine uptake than Edmonton and Calgary Health Zones. Some of the regions with the lowest coverage had few pharmacies offering the vaccine with minimal access to government-run clinics.(9); rural residents often had to travel to nonadjacent regions to access vaccination sites (9). In addition, rural residents have been shown to have lower levels of educations and may be more vaccine hesitant, which may contribute to lower coverage.(10–12) Consistent with a US-based study,(4) we found that coverage also varied between rural areas. The North zone had the lowest vaccination coverage, with only 55.8% having at least one dose. A US-based study found that disparities between rural counties was attributed to type of industry, with mining-dependent economies having lower vaccination coverage in comparison to recreational ones.(4)

The lower income neighborhoods reported lower vaccination coverage compared to the highest income neighborhoods. There was an apparent step-wise pattern in the crude vaccination coverage (Table 1), as well as in the multivariable model (Table 2). An Ontario-based study also showed that residents living in neighborhoods with high material deprivation had a low one-dose, two-dose, and booster dose coverage.(13) News reports have suggested that transportation, internet literacy, and language barriers are significant for low-income Albertans to receive vaccination.(10)

We hypothesized that areas with the highest COVID-19 risk quintile would have lower vaccination coverage. Similar work from Ontario showed that vaccination coverage was lower in areas defined as highest risk for COVID-19 infection.(11) Interestingly, in our study, we found that vaccination coverage was higher in areas defined as the highest COVID-19 risk quintile in comparison to the coverage in lowest risk quintile neighborhoods, with some regional variation. We found that most of the category 2 and 3 FSAs (highest COVID-19 risk neighborhood with high to medium coverage) lay in Northeast Calgary, and Northeast Edmonton. Although low vaccine turnouts occurred at the beginning of the pandemic in these areas and surrounding communities, there were focused efforts to improve vaccination by launching mobile clinics, targeted messaging, and engaging with community leaders, which might have improved vaccine uptake.(12) Additionally, Northeast Calgary (40%) and Northeast Edmonton have a high proportion of immigrant population; it is known that a significant proportion of immigrants are working as front-line workers, who may find social distancing difficult(14,15) We have previously shown that immigrants in Alberta have higher vaccination coverage than the non-immigrant population, which may also reflect this high risk employment.(16)

Furthermore, areas defined as high COVID-19 risk quintile and low coverage were in the Northwest of Calgary zone (Banff), Northwest of South zone (Brooks), and Northern Alberta (Fort McMurray, Fort McKay) (Figure 1). These areas have meat processing plants (Brooks),oil sands industry (Fort McMurray, Fort McKay), and recreational industry (Banff), which reported outbreaks at the initial stage of the pandemic.(17–19) Following the outbreaks, the infection might also have spread to nearby communities through workers and their families, which led to a high number of cases.(20) It was reported that these areas faced a vaccine supply issue at the initial stage of the pandemic, but were later targeted by improved vaccine delivery strategies. Notably, some of these areas (eg. Banff) currently have a high vaccination coverage (>80%).(21)

### Implications

These findings suggest that vaccine supply in rural areas needs to be timely and consistent. Focusing on largely rural health zones (North zone, South zone, and Central zone) and rural areas within other health zones may be an optimal strategy for vaccine promotion and delivery. Similarly, residents living in low income neighborhoods also require extra support. Furthermore, industrial workers who work in closed spaces with less chances of physical distancing need to be prioritized for vaccination.(22) Therefore, vaccine distribution and promotion strategies should not only look at individual risk factors but also to where people work and live. (23)

### Strengths and Limitations

This is one of the few studies conducted assessing the relationship of place-related variables with COVID-19 vaccine uptake. We used a population-based registry and immunization repository to assess vaccination status, making our findings representative of the entire province. We used multilevel analysis to account for the hierarchical nature of our data. However, our study has some limitations. We defined neighborhood risk quintiles based on the cumulative number of positive cases in each FSA. The high incidence may be the result of a big outbreak at the beginning of the pandemic and may not reflect the high-risk status in the later period. We could not include additional individual-level (e.g. educational status, political ideology, vaccine attitudes), household level variables (e.g. household size/income), and other place-based variables (e.g. distance to nearest clinic/pharmacy), due to lack of such data. Our estimated coverage is lower than coverage posted on the Alberta Health public website due to potential inclusion of people who departed the province without reporting to the health insurance program in the denominator.(24) We assumed that there is no systematic difference in departure pattern by place-based variables we have included in this analysis. If an individual received a vaccine after departing the province in other jurisdictions, they are misclassified as unvaccinated in the current analysis. Our analysis was aimed at assessing place-based distribution of vaccination and does not imply causality. Furthermore, the findings may not be generalizable to other jurisdictions.

## Conclusion

People living in rural areas, those in Central, South and North zones, those in the lowest income neighborhoods, and those in the lowest COVID–19 risk quintile neighborhoods had lower vaccine coverage at time of this study. Vaccine delivery policies and implementation strategies needs to focus on these areas to improve timely vaccination.

## Data Availability

The steward of the data used in this study is the Alberta Ministry of Health, who maintains the data for the purpose of health system administration. Thus, the authors are not at liberty to make the data publicly available

## Acknowledgements

We would like to thank Gary Gilham, Alberta Health for his support with creating map of Alberta showing neighborhood COVID-19 risk quintile and vaccination coverage. We would like to acknowledge Laura Reifferscheid, University of Alberta and Christa Smolarchuk, Alberta Health for their comments on the first draft of the manuscript.

## Contributions

SEM and YRP generated the idea for the paper, and YRP, CD and SEM prepared an analytical plan. YRP conducted data analyses. YRP prepared the first draft in consultation with SEM. YRP, CD, and SEM contributed to critical revision of the design and approved the final version. SEM supervised the research and is guarantor of the overall content.

## Ethical approval

The University of Alberta Health Research Ethics Board granted ethical approval for this study (Pro00114786).

