## Supplemental Figure 1 for "The influence of place on COVID-19 vaccine coverage in Alberta: A multilevel analysis"

Alberta population

(first quarter 2021 in population registry)

N=4,825,276

Living in Alberta during the study period

N=4,801,782

Exclude:

Left the province or died between Jan 2021 and August 31 2021,

n=23,494

Residents ≥12 years old included in analysis

N=3,945,103

Exclude:

First Nations residents, n=179,656

Age under 12 years, n=652,080

Population living in FSAs having <200 population, n=304

Lloydminster residents: n=24,374

Non-Alberta/missing postal code: n=252

Missing data on sex: n=13

Fig. A1 Sample selection flow chart
