## Supplemental Figure 2 for "The influence of place on COVID-19 vaccine coverage in Alberta: A multilevel analysis"

Numerator

Denominator

Population registry

Alberta population in the first quarter of 2021

Vital Statistics

Immunization data

Immunization and Adverse Reaction to Immunization (Imm/ARI) repository

Population after exclusion for death or out migration and age

First Nation’s registry

Eligible population for vaccination

Postal code Linking file

Fig. A2 Flow diagram showing database linkage
