## Supplemental Table 1 for "The influence of place on COVID-19 vaccine coverage in Alberta: A multilevel analysis"

**Table A1: Multilevel logistic regression analysis of factors associated with receipt of one dose of COVID-19 vaccine in Alberta, Canada ^a^**

|  | **Model 2**  **AOR (95% CI)** | **Model 3 AOR (95% CI)** | **Model 4 AOR (95% CI)** | **Model 5  AOR (95% CI)** |
| --- | --- | --- | --- | --- |
| **Age categories** | | | | |
| 12-17 years | Ref | Ref | Ref | Ref |
| 18-29 years | 1.00 (0.99-1.01) | 1.01 (1.0-1.02) | 1.00(0.94-1.06) | 1.01 (0.89-1.14) |
| 30-49 years | 1.05 (1.04-1.05) | 1.05 (1.04(1.06) | 1.03 (0.97-1.09) | 1.04 (0.92-1.18) |
| 50-64 years | 1.78 (1.77-1.79) | 1.79 (1.78-1.81) | 1.78 (1.67-1.88) | 1.77 (1.56-2.01) |
| 65 to 74 years | 2.68 (2.65-2.71) | 2.71 (2.68-2.74) | 2.59 (2.44-2.75) | 2.66 (2.35-3.02) |
| 75 years and above | 2.82 (2.79-2.86) | 2.89 (2.86-2.93) | 2.81(2.64-2.99) | 2.88 (2.54-3.26) |
| **Sex** | | | | |
| Male | Ref | Ref | Ref | Ref |
| Female | 1.21 (1.21-1.22) | 1.21 (1.21-1.22) | 1.21 (1.20-1.23) | 1.21 (1.20-1.23) |
| **Income Quintile** | | | | |
| Q1 (Lowest income) | N/A | Ref | Ref | Ref |
| Q2 | N/A | 1.14 (1.13-1.15) | 1.12 (1.07-1.17) | 1.22 (1.11-1.33) |
| Q3 | N/A | 1.28 (1.27-1.29) | 1.26 (1.21-1.32) | 1.37 (1.25-1.49) |
| Q4 | N/A | 1.42 (1.41-1.44) | 1.40 (1.34-1.46) | 1.50 (1.37-1.64) |
| Q5 (Highest income) | N/A | 1.67 (1.65-1.68) | 1.62 (1.55-1.70) | 1.76 (1.60-1.93) |
| **Geographic health zone** | | | | |
| Central zone | N/A | Ref | Ref | Ref |
| South zone | N/A | 0.93 (0.90-0.96) | 0.90 (0.86-0.94) | 1.17 (1.12 -1.22) |
| Calgary zone | N/A | 1.19 (1.16-1.23) | 1.23 (1.19-1.27) | 1.26 (1.21-1.31) |
| Edmonton zone | N/A | 1.03 (0.98-1.08) | 1.04 (0.98-1.10) | 1.11 (1.05-1.18) |
| North zone | N/A | 1.17 (1.12-1.23) | 1.02 (0.96-1.07) | 0.82 (0.77-0.86) |
| **Place of residence** | | | | |
| Rural | N/A | Ref | Ref | Ref |
| Metro | N/A | 1.28 (1.24-1.33) | 1.25 (1.20-1.30) | 1.37 (1.31-1.42) |
| Urban | N/A | 1.01 (0.98-1.03) | 1.00 (0.98-1.03) | 1.11 (1.08-1.14) |
| **Neighborhood COVID-19 risk level** | | | | |
| R5 (Lowest risk) | N/A | N/A | N/A | Ref |
| R1 (Highest risk) | N/A | N/A | N/A | 1.52 (1.12-2.05) |
| R2 (Higher risk) | N/A | N/A | N/A | 1.14 (0.93-1.39) |
| R3 (Medium risk) | N/A | N/A | N/A | 1.15 (0.96-1.38) |
| R4 (Lower risk) | N/A | N/A | N/A | 1.12 (0.96-1.32) |

*^a^ Notes:*

Model 1-Empty unconditional model with no exposure variables, not shown in the table

Model 2. Variables defining individual-level characteristics of the individuals (age categories and sex) added into the Model 1.

Model 3. Place-based variables defined at the postal code level (urban/rural place of residence, geographic health zone, neighborhood income quintile) were added into the Model 2.

Model 4. Variables showing significant association in the fixed effects in model 3 were included in the random statements into the Model 3.

Model 5 (final model): Place-based variable defined at the FSA level (Neighborhood COVID-19 risk level) was added to the Model 4.
